# Long period dynamics of viral load and antibodies for SARS-CoV-2 infection: an observational cohort study

**DOI:** 10.1101/2020.04.22.20071258

**Authors:** Jianping Huang, Tingting Mao, Shufei Li, Lianpeng Wu, Xueqin Xu, Huanzheng Li, Chenyang Xu, Feifei Su, Jianyi Dai, Jichan Shi, Jing Cai, Chongquan Huang, Xuan Lin, Dong Chen, XiaoLing Lin, Baochang Sun, Shaohua Tang

## Abstract

**OBJECTIVE:** To investigate the dynamics of viral RNA, IgM, and IgG and their relationships in patients with SARS-CoV-2 pneumonia over an 8-week period.

**DESIGN:** Retrospective, observational case series.

**SETTING:** Wenzhou Sixth People’s Hospital

**PARTICIPANTS:** Thirty-three patients with laboratory confirmed SARS-CoV-2 pneumonia admitted to hospital. Data were collected from January 27 to April 10, 2020.

**MAIN OUTCOME MEASURES:** Throat swabs, sputum, stool, and blood samples were collected, and viral load was measured by reverse transcription PCR (RT-PCR). Specific IgM and IgG against spike protein (S), spike protein receptor binding domain (RBD), and nucleocapsid (N) were analyzed.

**RESULTS:** At the early stages of symptom onset, SARS-CoV-2 viral load is higher in throat swabs and sputum, but lower in stool. The median (IQR) time of undetectable viral RNA in throat swab, sputum, and stool was 18.5 (13.25-22) days, 22 (18.5-27.5) days, and 17 (11.5-32) days, respectively. In sputum, 17 patients (51.5%) had undetectable viral RNA within 22 days (short persistence), and 16 (48.5%) had persistent viral RNA more than 22 days (long persistence). Three patients (9.1%) had a detectable relapse of viral RNA in sputum within two weeks of their discharge from the hospital. One patient had persistent viral RNA for 59 days or longer. The median (IQR) seroconversion time of anti-S IgM, anti-RBD IgM, and anti-N IgM was 10.5 (7.75-15.5) days, 14 (9-24) days, and 10 (7-14) days, respectively. The median (IQR) seroconversion time of anti-S IgG, anti-RBD IgG, and anti-N IgG was 10 (7.25-16.5) days, 13 (9-17) days, and 10 (7-14) days, respectively. By week 8 after symptom onset, IgM were negative in many of the previously positive patients, and IgG levels remained less than 50% of the peak levels in more than 20% of the patients. In about 40% of the patients, anti-RBD IgG levels were 4-times higher in convalescence than in acute phase. SARS-CoV-2 RNA coexisted with antibodies for more than 50 days. Anti-RBD IgM and IgG levels, including anti-RBD IgM levels at presentation and peak time, were significantly higher in viral RNA short persistence patients than in long persistence patients.

**CONCLUSION:** This study adds important new information about the features of viral load and antibody dynamics of SARS-CoV-2. It is clear from these results that the viral RNA persists in sputum and stool specimens for a relatively long time in many patients. Anti-RBD may also serve as a potential protective antibody against SARS-CoV-2 infection, as viral persistence appears to be related to anti-RBD levels. Earlier treatment intervention also appears to be a factor in viral persistence.

**WHAT IS ALREADY KNOWN ON THIS TOPIC:** There are several reports about the serum antibodies against SARS-CoV-2. However, most of them evaluate diagnostic accuracy. Only two articles report dynamics of SARS-CoV-2 viral RNA and antibodies with serial samples, but the observation periods are within 30 days. None of the studies investigate the profiles of SARS-CoV-2 viral load and antibodies in a long period. Three reports investigate profiles in respiratory samples, but there are no reports on the dynamics of the viral load in stool samples.

**WHAT THIS STUDY ADDS:** In both sputum and stool, SARS-CoV-2 RNA persists for a long time. The anti-RBD antibodies may involve in the clearance of SARS-CoV-2 infection. After eight weeks from symptom onset, IgM were negative in many of the previously positive patients, and IgG levels remained less than 50% of the peak levels in more than 20% of the patients. In about 40% of the patients, anti-RBD IgG levels increased 4-time higher in convalescence than in acute phase. Long persistence of SARS-CoV-2 viral RNA in sputum and stool presents challenges for management of the infection. The IgM/IgG comb test is better than single IgM test as a supplement diagnostic tool. Anti-RBD may be a protective antibody, and is valuable for development of vaccines.

## Introduction

A novel coronavirus, severe acute respiratory syndrome coronavirus 2 (SARS-CoV-2), has spread worldwide.^1-3^ As of April 18, 2020, more than 2,160,000 confirmed cases and more than 146,000 deaths have been reported. The clinical characteristics of SARS-CoV-2 pneumonia have been well defined. According to a large-scale epidemiological study in China, among the confirmed cases, 86.6% were aged between 30-79 years, 63.8% were men, 80.9% were considered as mild pneumonia, and 2.3% have died. ^4^ Common symptoms include fever, cough, fatigue, and lymphopenia etc. ^5-7^ On admission, half of the patients present typical radiological ground-glass opacity on chest computed tomography (CT). ^8^

Clearance of viral RNA and appearance of specific antibodies are essential for recovery from viral infection. SARS-CoV-2 viral load in respiratory samples rapidly increases soon after symptom onset and peaks at around 5-6 days. ^9-11^ During the convalescence period, the clearance of viral RNA in patients’ stool is delayed compared to oropharyngeal swabs. ^12^ As recommended by the National Health Commission of the People’s Republic of China, the criteria for discharge from the hospital are relieved symptoms and two successive negative viral nucleic acid results from respiratory samples separated by at least a 24 hour sampling interval. ^13^ It has been reported that some patients have had a relapse of the viral RNA after discharge from the hospital. ^14, 15^ However, this report included only four patients, and thus cannot show a complete picture of the patients discharged from hospitals. At present, reports about the profile of SARS-CoV-2 viral load have been mainly focused on relatively short periods (less than one month). The long-term dynamics of the viral load remain unclear.

Serological responses in patients are essential for understanding the immunological mechanisms and the recovery process of viral infection. It is also important for the development of serological diagnostic tools. Several assays have been developed for the detection of specific IgM and IgG against SARS-CoV-2. ^16-18^ These studies have focused either on the evaluation of detection methods, or the diagnostic value of IgM and IgG for SARS-CoV-2 infection. ^16, 17^ Only two studies investigated the dynamics of IgM and IgG. Those studies reported that the median seroconversion time of IgM and IgG was 12 and 14 days, respectively, and more patients had earlier seroconversion for IgG than IgM. ^18^ Due to the lack of serial results from the same cohort of patients over a longer period, the profile of specific IgM and IgG against SARS-CoV-2 remains unclear. Until now, antibody persistence after patients have been discharged from the hospital has been rarely reported, though this information is crucial for evaluating the immune profile of the recovered patients. Thus, the relationship between viral clearance and serological response has not been well investigated.

In this study, we investigated the profiles of viral RNA, IgM, and IgG in a group of patients with confirmed SARS-CoV-2 pneumonia over an 8-week period after symptom onset. Baseline characteristics and serial results of viral RNA in throat swabs, sputum, and stool specimens were monitored. Specific IgM and IgG against spike protein (S), spike protein receptor binding domain (RBD), and nucleocapsid (N) were also measured. In this way, the long-term relationship between viral RNA persistence and the dynamics of SARS-CoV-2 antibodies have been more clearly established.

## Methods

### Study design and participants

This retrospective study was completed in Wenzhou Sixth People’s Hospital, Wenzhou Central Hospital Group, one of the designated hospitals to treat patients with SARS-CoV-2 pneumonia. A total of 33 patients admitted to Wenzhou Sixth People’s Hospital, Wenzhou Central Hospital Group, from January 27 to February 13, 2020 were included in this study. Among the patients, 31 (93.9%) were moderate, and two were severe SARS-CoV-2 pneumonia according to interim guidance issued by the WHO ^19^ and National Health Commission of the People’s Republic of China. ^13^ The patients were followed up for 12-38 days (median, 30 days) after discharge from the hospital.

This study was approved by the Ethics Commission of Wenzhou Central Hospital (L2020-01-036). Patient Consent Form was signed by patients themselves.

### Data collection

A team of trained physicians and medical students reviewed all available electronic medical records, radiological findings, and laboratory examinations for all hospitalized patients with confirmed SARS-CoV-2 infection. Data were recorded on a standardized data collection form. Demographic data, symptom onset time, clinical features, radiological findings, routine laboratory results, and the results of SARS-CoV-2 viral RNA in throat swabs, sputum, and stool samples were recorded during hospitalization and follow-up. Results of specific IgM and IgG antibodies against SARS-CoV-2 S, RBD, and N were measured during hospitalization and follow-up. Any missing or uncertain records were clarified through communication with the physicians responsible for the patients in question, or with the patients themselves.

### Statistical analysis

Continuous variables were presented as medians with interquartile ranges (IQRs) and means with standard deviations (SD). For categorical variables, percentages of patients in each category were analyzed. Differences between patients with sputum viral RNA short-persistence (viral RNA undetectable within 22 days) and those with long-persistence (viral RNA persists more than 22 days) were assessed by two sample *t* test or the Wilcoxon ranks sum test, depending on parametric or nonparametric data for continuous variables. Spearman’s correlation analysis was used to assess relationships between parameters. All analyses were done with SPSS version 24.0 (IBM, Armonk, NY, USA). Tests were two-sided with significance assessed at the 0.05 level.

## Results

A total of 33 patients admitted to Wenzhou Sixth People’s Hospital, Wenzhou Central Hospital Group, from January 27 to February 13, 2020, were included in this study. The median age of the patients was 47 years old (range, 2-84) and 29 (87.9%) of them were less than 60 years old. Among the patients, 17 (51.5%) were men, and 12 (36.4%) had comorbid illness (Supplemental Table 1)

The most common symptoms were fever (19, 57.6%), cough (17, 51.5%), expectoration (4, 12.1%), fatigue (3, 9.1%), and diarrhea (3, 9.1%). Eight patients (24.2%) had only a single symptom, and six patients (18.2%) were asymptomatic. Twenty-four patients (72.7%) had respiratory symptoms only, two (6.1%) had symptoms in both the respiratory and gastrointestinal tracts, and one (3.0%) had gastrointestinal symptoms only. Chest CT confirmed that 27 patients (81.8%) had bilateral infiltrates, and six (18.2%) had unilateral infiltrates. The median time period from symptom onset to hospital admission was 3 days (range, 0.5-15; IQR 1.5-7), and the median time of hospitalization was 22 days (range, 16-30; IQR, 18-24). The median (IQR) follow-up time after the patients’ discharge from the hospital was 30 days (range, 12-38; IQR, 25-33.5).

Routine laboratory parameters at admission and the first return visit time were available for analysis. Most of the laboratory parameters at admission were normal in all of the patients. Abnormal parameters observed in the patients at admission were abnormal white blood cell count (9, 27.3%), neutrophil count (4, 12.1%), lymphocyte count (5, 15.2%), platelet count (1, 3.0%), C-reactive protein (CRP) (15, 45.5%), creatinine kinase (CK) (2, 6.1%), lactate dehydrogenase (LDH) (10, 30.3%), blood urea nitrogen (BUN) (9, 27.3%), and blood creatinine (Cr) (8, 24.2%). Almost all of the patients had normal laboratory parameters upon their return visit one week after discharge (Supplemental Table 2).

All of the patients received antiviral treatment during hospitalization. Atomized interferon, lopinavir and ritonavir, arbidol, ribavirin, and Lianhuaqingwen (Chinese traditional medicine) were used in combination. Atomized interferon was used in all patients. Lopinavir and ritonavir was used in 32 (97.0%) of the patients. Lianhuaqingwen, which was believed to relieve fever and cough, was used in 28 (84.8%) of patients. Arbidol was used in 16 (48.5%) of the patients. All patients recovered well. The most common side effects were elevation of triglycerides (16, 48.5%), elevated transaminases (10, 30.3%) and diarrhea (9, 27.3%).

A total of 263 respiratory samples and 135 stool samples were obtained from 33 patients. SARS-CoV-2 viral RNA was detected by real-time reverse transcription PCR (RT-PCR). The cross time (Ct) value reflects viral load in the sample. Since the maximum Ct value was 40 in our study, we use L = 40-Ct to indicate the viral load in samples. At the earliest stages of symptom onset, SARS-CoV-2 viral load is higher in throat swabs and sputum, but lower in stool. Viral load in throat swabs declined quickly, and it became undetectable among most patients by three weeks after symptom onset. In sputum, viral load declined slower, and most patients were undetectable by week 5 after onset. Viral load in stool declined steadily, and many patients had persistent viral RNA for more than five weeks (Figure 1). The median (IQR) time of undetectable viral RNA in throat swab, sputum, and stool was 18.5 (13.25-22) days, 22 (18.5-27.5) days, and 17 (11.5-32) days, respectively. Compared to throat swabs, viral loads in sputum and stool declined significantly slower (vs stool, *P*=0.046; vs sputum, *P*=0.005). However, the decline of viral loads in sputum and stool showed no significant difference (*P*=0.391). Since the median time of undetectable viral RNA in sputum was 22 days, we divided the patients into two groups: viral RNA short-persistence (viral RNA undetectable within 22 days) and viral RNA long-persistence (viral RNA persists more than 22 days). In sputum specimens, 17 patients (51.5%) had undetectable viral RNA within 22 days, and 16 patients (48.5%) had persistent viral RNA for more than 22 days. Three patients (9.1%) showed a detectable relapse of viral RNA in sputum when they were tested two weeks after discharge from the hospital. One patient had persistent viral RNA for 59 days or longer. SARS-CoV-2 viral loads at the first detection after onset were not corelated with age in throat swabs (Spearman’s *P*=0.836), sputum (Spearman’s *P*=0.459) or stool specimens (Spearman’s *P*=0.581). Hospitalization period, which was determined by two continuously undetectable viral RNA results in respiratory specimens, was not corelated with age (Spearman’s *P* =0.753).

**Figure 1.**
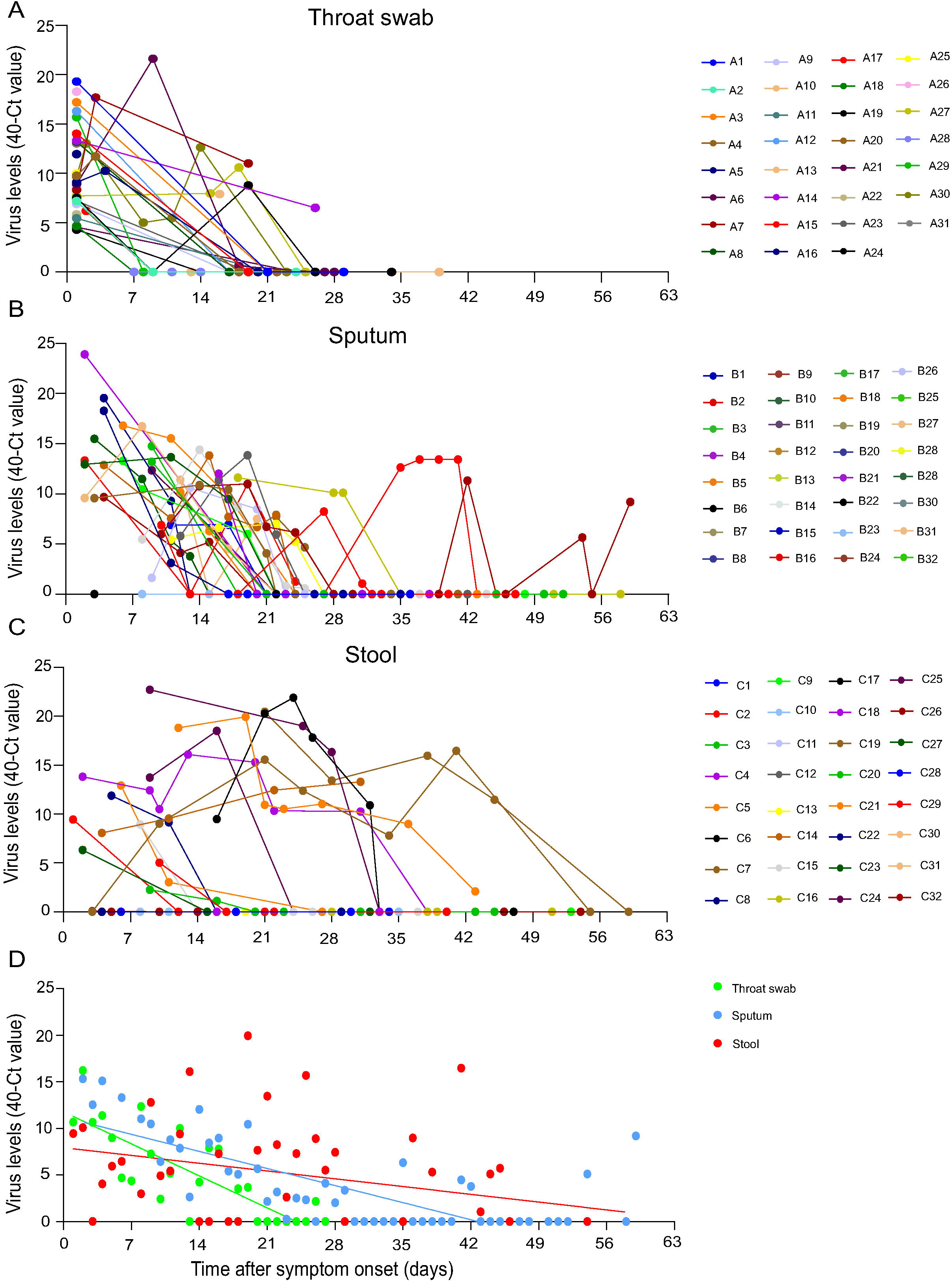
Viral dynamics of SARS-CoV-2 in all patients (n=33) Each line represents an individual patient. Viral dynamics in throat swab (A), sputum (B), and stool (C). A comparison of viral loads across different types of specimens is also shown (D).

A total of 171 serum samples were available for the analysis of SARS-CoV-2 S, RBD, and N specific IgM and IgG antibodies. All of the antibodies were tested by a chemiluminescence assay (Nanjing RealMind Biotech Co., Ltd, Nanjing, China). The levels of the antibodies were expressed as arbitrary unit (AU) per microliter (mL), which was defined by the manufacturer. The median (IQR) seroconversion time of anti-S IgM, anti-RBD IgM, and anti-N IgM was 10.5 (7.75-15.5), 14 (9-24), and 10 (7-14) days, respectively. The median (IQR) seroconversion time of anti-S IgG, anti-RBD IgG, and anti-N IgG were 10 (7.25-16.5), 13 (9-17), and 10 (7-14) days, respectively. Anti-RBD antibodies appeared later than anti-S (anti-RBD IgM vs. anti-S IgM, *P*=0.007; anti-RBD IgG vs. anti-S IgG, *P*=0.006). Anti-RBD antibodies appeared later than anti-N, but the difference was not statistically significant (anti-RBD IgM vs. anti-N IgM, *P*=0.054; anti-RBD IgG vs. anti-N IgG, *P*=0.078). For IgM and IgG seroconversion, IgG was earlier than IgM in many patients for S (4 [12.1%] of 33), RBD (9 [27.3%] of 33), and N (3 [9.1%] of 33), respectively. There was only one patient who had earlier anti-RBD IgM than IgG. Most of the patients showed simultaneous seroconversion of IgM and IgG. When comparing the seropositivity of IgM and IgG across the entire study period, 6 (18.2%), 9 (27.3%), and 14 (42.4%) out of 33 patients had negative IgM of anti-S, anti-RBD, and anti-N, respectively. Three patients (9.1%) were negative for all three IgM antibodies. One patient (3.0%) had negative anti-N IgG, and another patient (3.0%) had negative anti-RBD IgG. All 33 patients (100%) had positive anti-S IgG. For IgM, the median (IQR) peak time of anti-S, anti-RBD, and anti-N was 22 (15.75-32), 25 (18-38), and 14 (9-23) days after symptom onset, respectively. For IgG, the median (IQR) peak time of anti-S, anti-RBD, and anti-N was 30.5 (20.5-39), 30 (22-39), and 28 (17-37) days after symptom onset, respectively. Seropositivity of IgM declined quickly, and by week 8 after symptom onset, IgM were negative in many of the previously positive patients (anti-S, 9 of 27 [33.3%]; anti-RBD, 5 of 24 [20.8%]; anti-N, 12 of 19 [63.2%]). In contrast, IgG remained positive after seroconversion, but the antibody levels still experienced an overall decline, and in more than 20% of the patients (anti-S, 4 [12.1%]; anti-RBD, 7 [21.2%]; anti-N, 7 [21.2%]), IgG levels remained less than 50% of the peak levels (Figures 2, 3, and 4). When comparing the IgG levels between the acute and convalescent phases, a 4-fold increase was observed in 6 patients (18.2%) for anti-S, 12 patients (36.4%) for anti-RBD, and 3 patients (9.1%) for anti-N, respectively.

**Figure 2.**
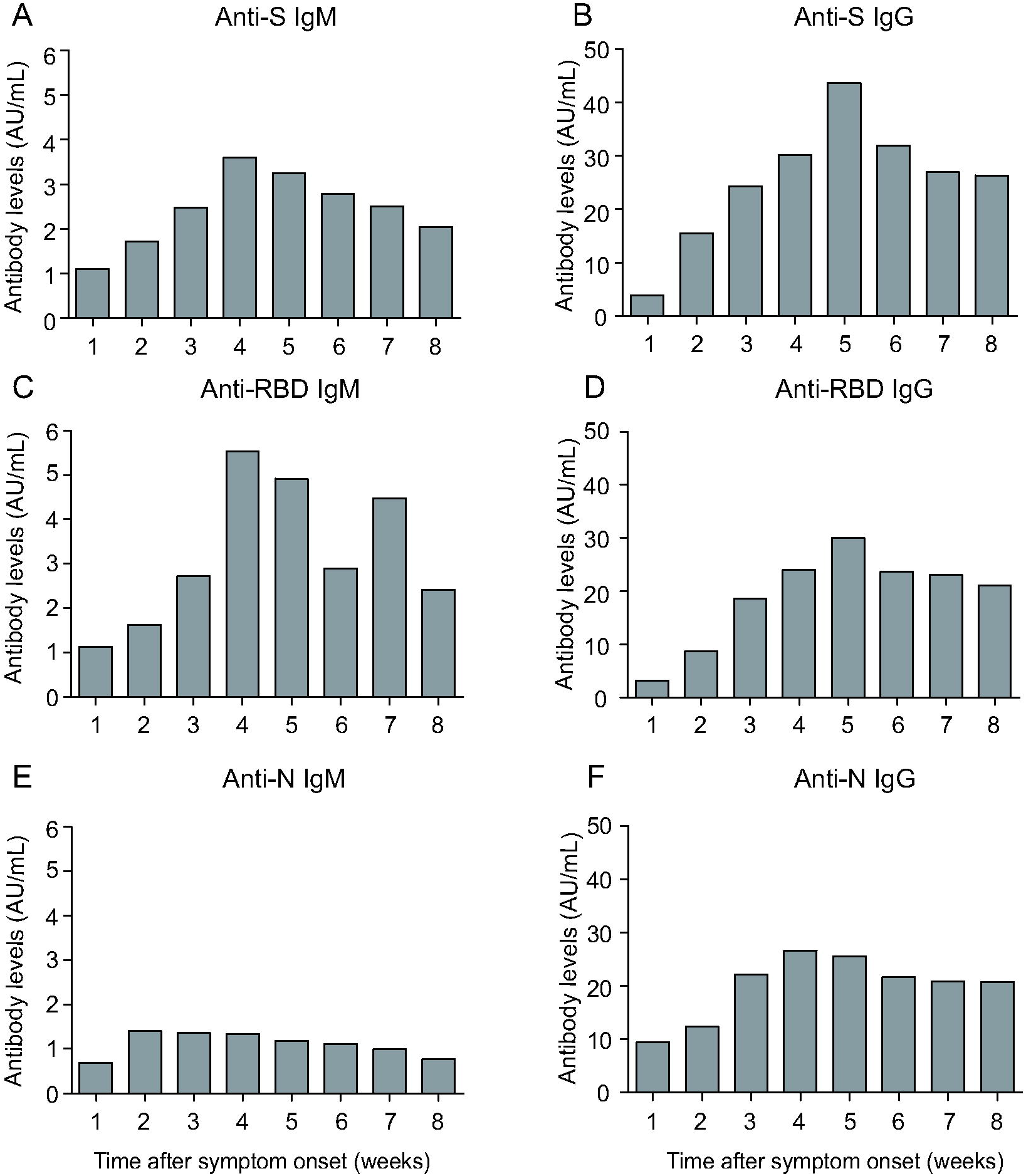
Dynamics of antibodies against SARS-CoV-2 in patients. Each column represents the mean levels of the antibody in one week.

**Figure 3.**
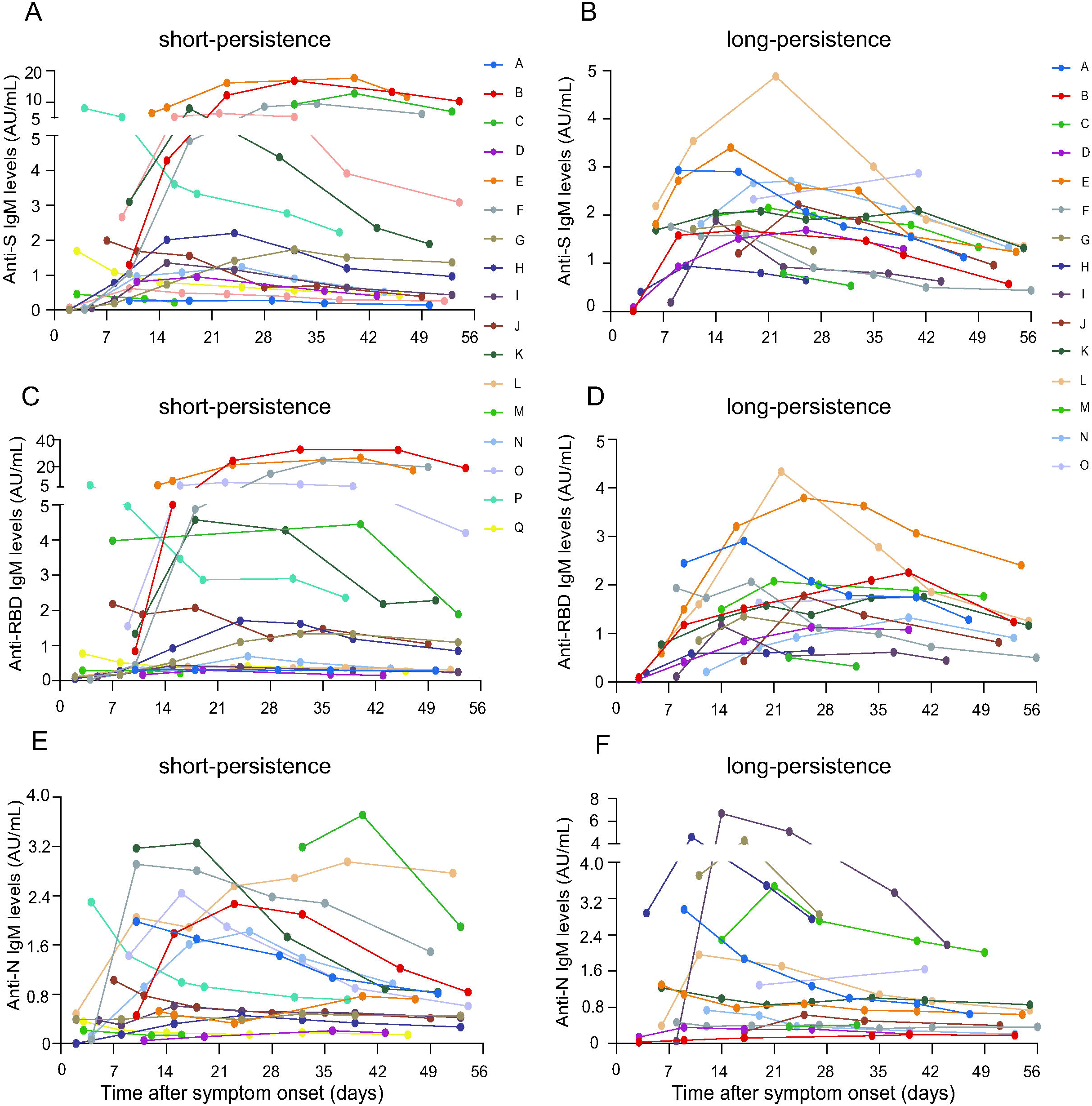
Comparing the profiles of serum anti-S, anti-RBD, and anti-N IgM between the SARS-CoV-2 RNA short and long persistence groups. Each line represents an individual patient. S = spike protein. RBD = spike protein receptor binding domain. N = nucleocapsid. One patient in the long persistence group had very high immunoglobulin levels due to the presence of a tumor and was removed from the figure.

**Figure 4.**
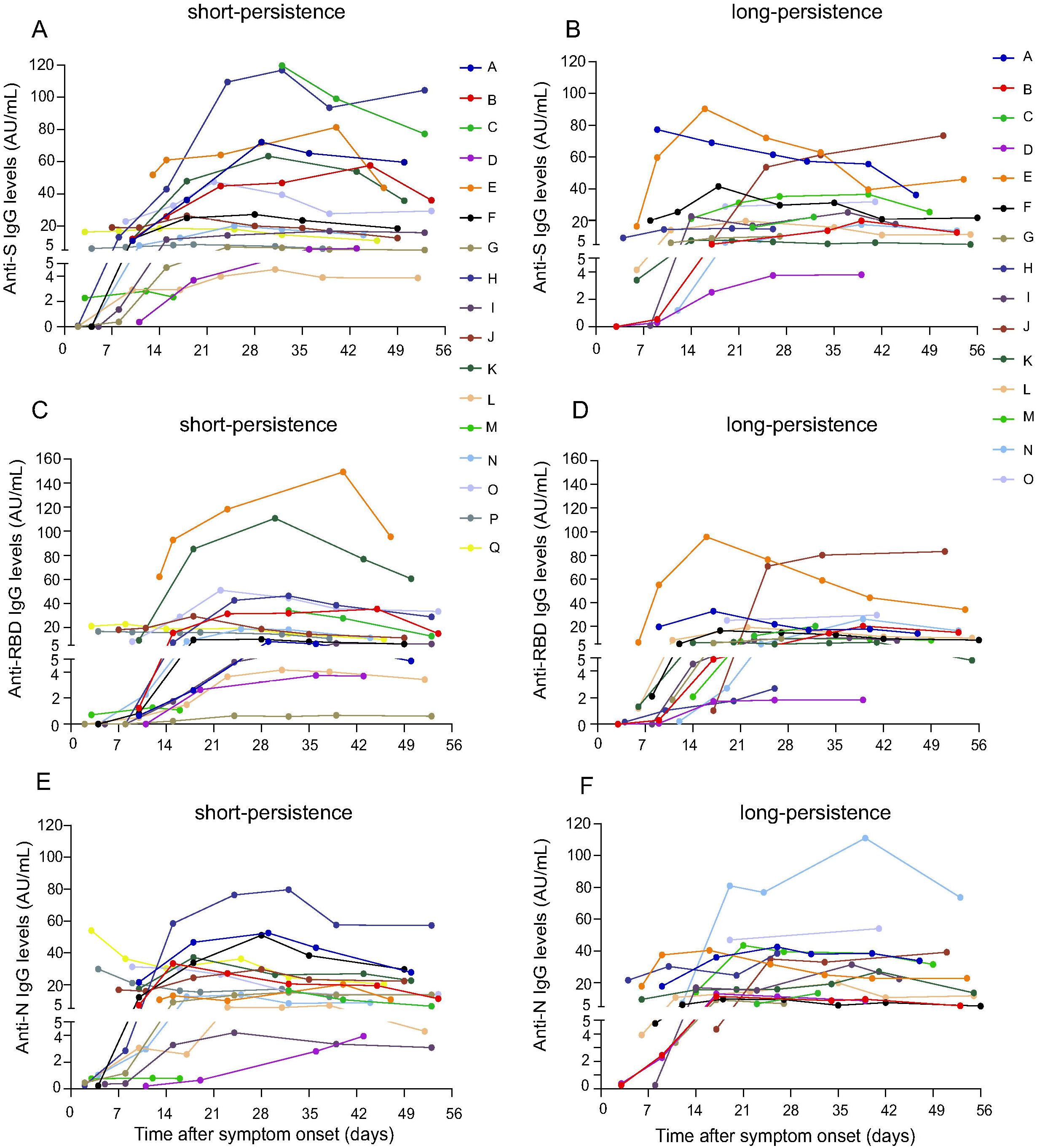
Comparing profiles of serum anti-S, anti-RBD, and anti-N IgG between the SARS-CoV-2 RNA short and long persistence groups. Each line represents an individual patient. S = spike protein. RBD = spike protein receptor binding domain. N = nucleocapsid. One patient in the long persistence group had very high immunoglobulin levels due to the presence of a tumor and was removed from the figure.

Since detection of SARS-CoV-2 RNA in sputum is a routine test during treatment, and is commonly used in clinical settings, the relationship between the persistence of sputum viral RNA and antibodies was analyzed (Figures 3, 4, and 5). Compared to the viral RNA long persistence group, patients in the short persistence group had higher anti-S Ig G (*P*=0.000), anti-RBD IgM (*P*=0.000), and anti-RBD IgG levels (*P*=0.043). Anti-RBD IgM levels at seroconversion (*P*=0.019) and the peak level of anti-RBD IgM (*P*=0.037) were also significantly higher in the short persistence group. It was also observed that the time from symptom onset to hospital admission was significantly different between the short persistence and long persistence groups (*P*=0.048).

**Figure 5.**
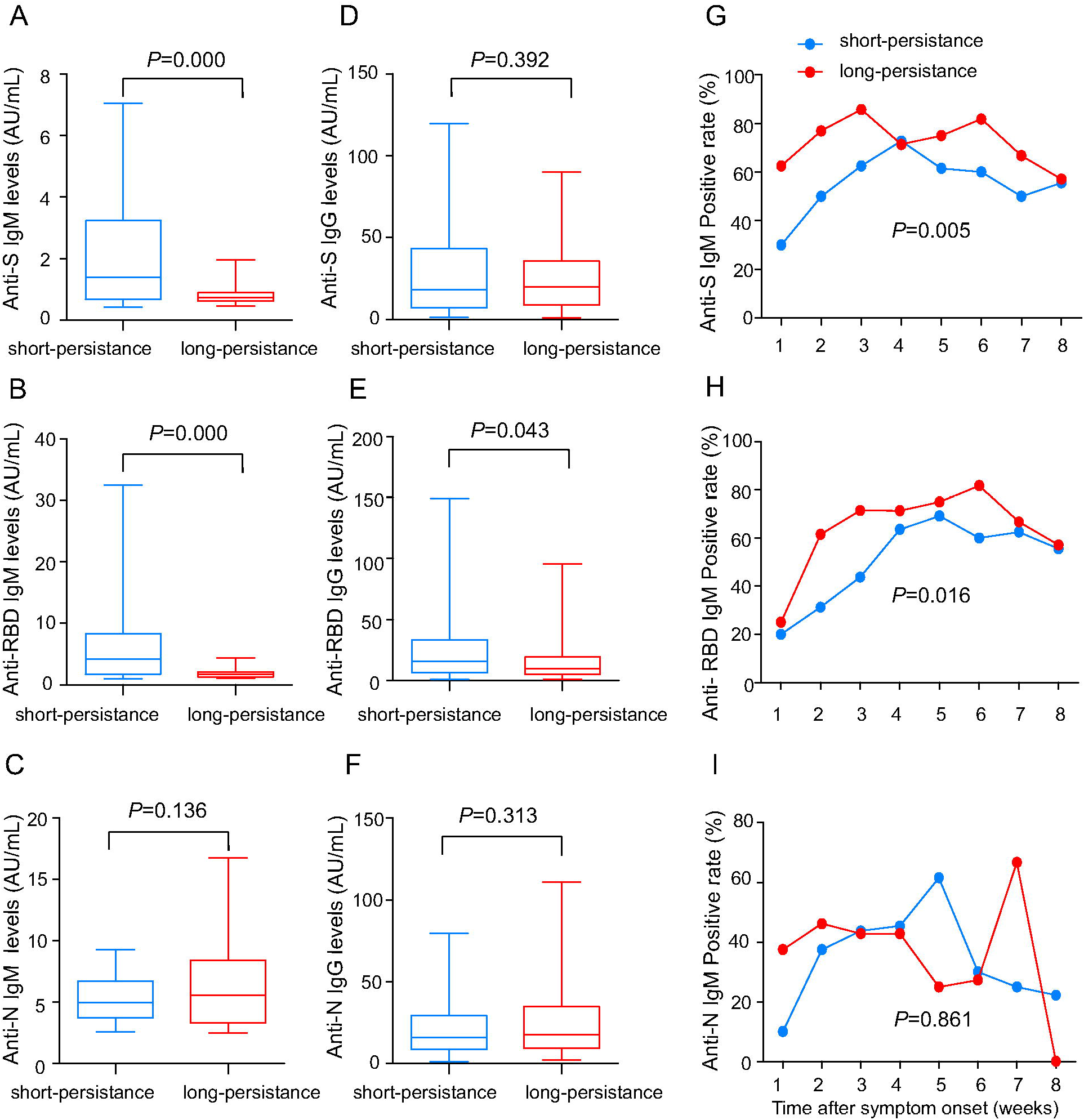
Comparison of levels and seropositivity of antibodies against SARS-CoV-2 between viral RNA short and long persistence groups. S = spike protein. RBD = spike protein receptor binding domain. N = nucleocapsid. Only seropositive results (>1 AU/mL) were included in the figure. One patient in the long persistence group had very high immunoglobulin levels due to the presence of a tumor and was removed from the figure.

## Discussion

We investigated the serial viral load and dynamics of antibodies from patients infected with SARS-CoV-2 over an eight-week period following the onset of symptoms. In throat swab specimens, the SARS-CoV-2 viral load was high at the beginning of infection but declined quickly. The viral load in stool samples was low in the initial period but declined slowly. In both sputum and stool, viral RNA persisted for a long time. Simultaneous seroconversion of IgM and IgG was observed in most of the patients. IgM peaked in the fourth week (except for anti-N IgM, which peaked in the second week), and IgG peaked between the fourth and fifth week after the onset of symptoms. High anti-RBD antibody levels were associated with short persistence of SARS-CoV-2 RNA in sputum.

SARS-CoV-2 viral load varies across different types of specimens. ^20^ In this study, we observed that in throat swab samples, the viral load was high in the early stages of infection, but it became undetectable by three weeks following symptom onset. Pan et al previously reported the viral load in several types of specimens from two patients ^10^, and To and colleagues have reported the viral load in posterior oropharyngeal saliva samples. ^11^ Combining those studies with our own results here, we can conclude that in upper respiratory samples, SARS-CoV-2 viral load is high shortly after symptom onset but declines quickly.

The detection of SARS-CoV-2 RNA in respiratory specimens is included in the guidelines for monitoring treatment. According to these guidelines, patients will be discharged from hospitals if they have experienced relief of symptoms and had two successive negative viral RNA tests from respiratory specimens across at least a 24 h sampling interval. ^13^ We observed that in sputum, viral load declined with time, and most patients were undetectable by week 5 after the onset of symptoms. However, positive viral RNA was detected in about 10% of our patients two weeks after their discharge from the hospital. In addition, one patient in our cohort had positive viral RNA for 59 days after symptom onset and would go on to persist even longer. This means that SARS-CoV-2 may persist for a long time in some patients. Whether “relapsed patients” are truly relapsed, or the result of false negative viral RNA tests, needs to be explored further.

Our study also investigated SARS-CoV-2 viral load in serial stool specimens. The viral load in stool was lower than what was observed in throat swabs and sputum at the beginning of infection but declined slowly. Many patients had persistent viral RNA in stool for more than five weeks. Since fluctuations of viral RNA in respiratory specimens were observed in most of the patients, false negative results will be inevitable in such specimens. Detection of viral RNA in stool specimens may thus act as a useful complement for monitoring treatments.

Currently, serological tests are widely used for diagnosis, and SARS-CoV-2 IgM or IgM/IgG tests have been approved under emergency use authorization (EUA) in many countries. In most of the known viral infections, seroconversion of IgM occurs earlier than seroconversion of IgG. ^21-24^ Hence, specific IgM is often used as a diagnostic marker for acute viral infection. Our results indicate that in SARS-CoV-2 infected patients, simultaneous seroconversion of IgM and IgG was observed in 75% of the patients. Almost no patient had earlier seroconversion of IgM than IgG. Nearly 10% of the patients had negative IgM across the entire study period. To and colleagues also reported that in most of SARS-CoV-2 infected patients, seroconversion of IgG was earlier than IgM, and they thought that this might be due to low sensitivity of their IgM test. ^11^ Combining the results together, it appears that a single IgM test may not be a reliable index for an auxiliary diagnosis of SARS-CoV-2 infection. Convalescent IgM/IgG testing will likely be better than single IgM for patients for whom nucleic acid tests are not available, or those who have typical SARS-CoV-2 symptoms but are negative for nucleic acid detection.

The coexistence of SARS-CoV-2 viral RNA and specific antibodies has been reported by several studies, ^11, 17, 18^ and has been observed in patients with severe acute respiratory syndrome (SARS) and middle eastern respiratory syndrome (MERS). ^25, 26^ Our results shown that in some patients, SARS-CoV-2 RNA coexisted with antibodies for more than 50 days. These results raise concerns about the existence of protective antibodies against SARS-CoV-2, since one of the key roles of protective antibodies is to clear viral infection. ^27^ To and colleagues reported that anti-SARS-CoV-2-N or anti-SARS-CoV-2-RBD IgG levels correlated with virus neutralization titre. ^11^ In this study, we found that anti-SARS-CoV-2 IgG peaked within about 30 days after symptom onset. In about 36% of the patients, anti-RBD IgG increased more than four-fold within one to two weeks after seroconversion and persisted for more than four weeks. We also found that anti-RBD IgM and IgG levels, including anti-RBD IgM levels at presentation and peak time, were significantly higher in patients showing viral RNA short persistence than in patients showing long persistence. These data imply that the anti-RBD antibodies may be involved in the clearance of SARS-CoV-2 infection and would be potential candidates for a protective antibody. Further studies are needed to identify whether RBD is a useful epitope for developing vaccines against SARS-CoV-2.

We also observed that the length of time from symptom onset to hospital admission was associated with SARS-CoV-2 viral RNA clearance. In addition, the seropositive rate for anti-S and anti-RBD IgM was significantly higher in viral RNA long persistence patients. Based on current evidence, the relationship between late treatment and seropositivity for anti-S or anti-RBD IgM is undetermined. It is possible that the higher seropositive rate is due to a longer infection time and delayed treatment.

However, our data imply that shorter delay is linked to better recovery. “Early identification, early hospitalization” is thus likely to be an efficient strategy for the prevention and management of SARS-CoV-2 infection. Our study has some limitations. First, the patient number in our study is relatively small, and only 33 patients were included in this study. This is, of course, a common weakness of studies of emerging infectious diseases. Second, samples for viral RNA and antibody detection were collected weekly and not on a daily basis. It was also difficult to collect samples at exactly one-week intervals. In some patients, initial samples were even missed due to the emerging situation for SARS-CoV-2 infection.

In conclusion, this study adds important new information about the features of viral load and antibody dynamics of SARS-CoV-2. It is clear from these results that the viral RNA persists in sputum and stool specimens for a relatively long time in many patients. Anti-RBD may also serve as a potential protective antibody against SARS-CoV-2 infection, as viral persistence appears to be related to anti-RBD levels. Earlier treatment intervention also appears to be a factor in viral persistence. Further studies are needed to better understand the virological and immunological features of SARS-CoV-2 infection, which are important for the prevention, treatment and control of the spread of this disease.

## Data Availability

All data included in this study are available upon request by contact with the corresponding author.

https://mail.163.com/js6/main.jsp?sid=bAZugYCSydNlElSYogSShrpZfhqgCrxI&df=mail163_letter#module=welcome.WelcomeModule%7C%7B%7D

## Footnotes

### Contributors

SHT designed the study, and had full access to all data in the study. JPH and TTM wrote the report and contributed equally to this paper. SHT and TTM analyzed the data. JPH, FFS, JYD, JCS and JC contributed to patient recruitment, data collection and provided clinical supervision. SFL, LPW, XQX, HZL and CYX conducted the experiments and collected the data. TTM and XL made the figures and tables. DC performed the literature search. CQH and BCS had a role in data collection.

### Ethical approval

This study was approved by the Ethics Commission of Wenzhou Central Hospital (L2020-01-036).

### Patient consent

Written informed consent was waived due to the emergency state of the disease.

### Data sharing

No additional data available.

### Transparency declaration

The lead author and the manuscript’s guarantor affirms that the manuscript is an honest, accurate, and transparent account of the study being reported; that no important aspects of the study have been omitted; and that any discrepancies from the study as planned (and, if relevant, registered) have been explained.

